# Evaluation and Treatment of Severe SARS-CoV-2 Pneumonia: A Scoping Review

**DOI:** 10.1101/2023.06.13.23291341

**Authors:** Xiyue Li, Jianbo Huang

## Abstract

**Purpose:** Severe SARS-CoV-2 pneumonia remains incompletely understood. We aimed to summarize current evidence regarding clinical features, laboratory findings, and treatment of severe pneumonia.

**Methods:** Online databases were searched from December 1, 2019, to April 15, 2020 related to SARS-CoV-2. The titles and abstracts in English or Chinese for articles were screened. Studies containing more than 10 adult patients with severe pneumonia and presenting data on clinical features and laboratory findings were selected and extracted independently by two reviewers.

**Results:** We identified a total of 13 articles including one from Italy representing the majority of cases, and the remainder from China. Over the 2,129 severe pneumonia in these 13 articles, the mean ages ranged from 49 to 64 years. Patients typically presented with hypertension as the most common comorbid factor, fever as the most common symptom, and acute respiratory distress syndrome as the most common complication. As compared to non-severe pneumonia, severe pneumonia featured lower counts of lymphocytes, CD8+ and CD4+ T cells, and higher levels of D-dimer, lactate dehydrogenase, IL-6 and IL-10. There is a lack of evidence for using antivirals, and a debate on using corticosteroids in treatment.

**Conclusions:** This is the first systematic summarization of the aspects of severe pneumonia. Older age, comorbidities, laboratory findings might be the predisposing factors of disease severity. Multicenter- and large population-designed studies, with confounding controlled and long enough to accommodate follow-ups, are urgently required to provide the guidance to disease management.

## Introduction

Outbreak of a novel coronavirus disease, COVID-19, initiated in Wuhan, China in December of 2019. This disease is caused by the severe acute respiratory syndrome coronavirus 2, SARS-CoV-2. Coronaviruses are a large family of viruses that cause illnesses ranging from the common cold to more severe diseases such as Middle East Respiratory Syndrome (MERS) and Severe Acute Respiratory Syndrome (SARS). These diseases are primarily transmitted by respiratory droplets and close personal contact [1]. As of June 4, 2020, there were 6,475,644 laboratory-confirmed cases have been reported, including 386,544 deaths globally [2].

Cases with severe SARS-CoV-2 pneumonia (Referred to as severe pneumonia) possessed the high case-fatality [3, 4] and required intense attention [5] according to previous studies. A complete understanding of severe pneumonia should facilitate faster diagnosis and more effective treatment. Unfortunately, information about the characteristics and treatments of severe pneumonia is limited, heterogeneous, and not consistently defined. This scoping review aimed to summarize current evidence regarding the demographics, comorbidities, clinical manifestations, laboratory findings, therapies, and outcomes of severe pneumonia.

## Methods

The protocol for this scoping review is provided in the supplementary materials. The review was conducted according to the framework described by Arksey and O’Malley [6], and was in accordance with the Preferred Reporting Items for Systematic Reviews and Meta-Analyses extension for scoping reviews (PRISMA-ScR) statement [7]. After identifying relevant studies that met the selection and inclusion criteria, data was examined and tabulated. Findings from this data were used to draw conclusions according to our research aims. All findings and statements regarding the severe pneumonia in this scoping review were based on the information from the included studies.

### Identifying relevant studies

To include the maximum number of relevant articles pertaining to severe pneumonia, we searched for the terms “2019 novel coronavirus”, “2019-nCoV”, “Coronavirus disease 2019 virus”, “COVID-19 virus”, “SARS-CoV-2” and “SARS2”. These terms were searched in the title and/or abstract field of literature published between December 1, 2019 and April 15, 2020 in PubMed, Medline, Embase, Web of Science, preprint servers for health sciences (MedRxiv and BioRxiv), Cochrane library and China National Knowledge Infrastructure databases. Duplicates were removed using EndNote X8 software. We did not set a language restriction in the process of literature search, the final search strategy for PubMed can be found in the supplementary methods. We manually reviewed titles and abstracts in English or Chinese, regardless of the language of the publication. For publications in languages beyond English and Chinese, none were proceeded for full-text screening after reviewing the publications’ abstract.

### Study selection and inclusion criteria

Potential articles were reviewed to ensure that they presented clinical features or laboratory findings of patients with severe pneumonia. Two authors (X.L. and H.Z.) independently examined the titles, abstracts, and full texts of publications to determine if they would be included in the study. Disagreement was resolved either through discussion with each other, or with a third researcher (Z.C.) vote if necessary. The inclusion criteria contained: (1) Adult patients with laboratory-confirmed SARS-CoV-2 pneumonia: positive result of real-time reverse transcriptase–polymerase chain reaction (RT-PCR) assay of nasal or pharyngeal swabs; (2) Severe pneumonia with at least one of the following: i) respiratory rate ≥ 30 breaths/min, ii) SpO_2_ ≤ 93% on room air, iii) PaO_2_/FiO_2_ ≤ 300 mmHg [8]; (3) More than 10 patients with severe pneumonia included in the study; (4) Any clinical and/or laboratory data presented. The PaO_2_/FiO_2_ in acute respiratory distress syndrome (ARDS) is at least less than 300 mmHg based on the Berlin definition [9]. Any type of study design such as single center- or multicenter-based, small or large sample size-based, retrospective or prospective, observational, and randomized controlled trial (RCT), would be selected if the study met the inclusion criteria.

### Data charting and quality assessment

The charted information included the name of the first author, study institution, study type, definition of severe pneumonia, case number, gender, age, comorbidities, symptoms, complications, laboratory findings, treatment, prognosis, and the comparison between severe and non-severe pneumonia when appropriate. Some data about cytokines and lymphocyte subsets were extracted from graphs with software GetData Graph Digitizer (version 2.26). Data was inputted into an excel spreadsheet with all data charted first independently by two authors (X.L. and H.Z.) and then compared. Disagreement on charted information was resolved either through a discussion between each other, or with a third researcher (Z.C.) vote if necessary.

With the Joanna Briggs Institute (JBI) critical appraisal tool for case series, or cross-sectional studies if comparing severe and non-severe pneumonia [10], two researchers (X.L. and H.Z.) independently assessed the methodological quality of individual study and determined the extent to which the study has addressed the possibility of bias in its design, conduct and analysis. The overall appraisal on each source of evidence was decided to include or exclude the study. Disagreement on quality assessment was resolved either through a discussion between each other, or with a third researcher (Z.C.) vote if necessary.

### Summarizing the findings

Articles were classified into one of the following research domains: demographics, comorbidities, clinical manifestations, complications, laboratory findings that including routine tests, cytokines and counts of lymphocyte subsets, treatments, and prognosis. We counted the number of studies included and summarized the findings. We calculated overall ratio of each item in comorbidities, symptoms and complications, and narratively described overall ratio with the range of lowest and highest values amongst the related studies. Categorical data were expressed as numbers (%). Continuous data were expressed as medians and interquartile ranges (IQR) or means and standard deviations. The significant differences between severe and non-severe pneumonia were recorded if appropriate.

## Results

The literature search identified 4,824 articles. After discarding 1,883 duplicates, the titles and abstracts of 2,941 articles were screened. Sixty-three articles were retrieved for full-text evaluation. Of these, 50 articles were excluded for the following reasons: the study did not define severe pneumonia (n=25); clinical or laboratory data were unavailable (n=13); the study possessed less than 10 severe pneumonia (n=12). Thirteen studies (12 in English, one in Chinese [11]) with 2,129 cases of severe pneumonia meeting our inclusion criteria were included [3, 11–22], one from Italy with by far the majority of the severe pneumonia; and the remainder from China (Figure 1).

**Figure 1.**
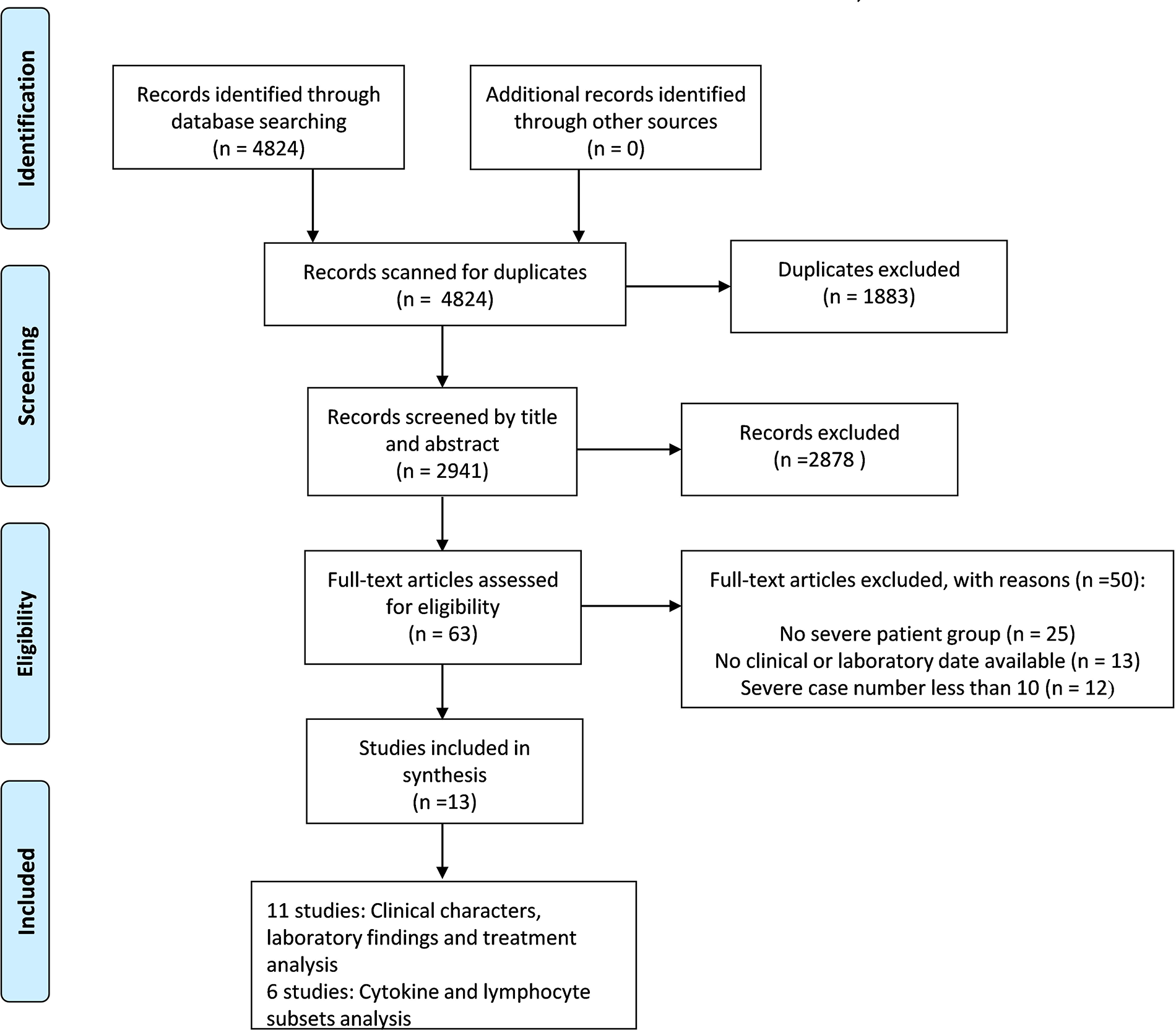
Literature search results for identification, exclusion, and selection of studies. The literature search identified 4,824 articles. After discarding 1,883 duplicates, the titles and abstracts of 2,941 articles were screened. 63 articles were retrieved for full-text evaluation. Thirteen articles met our inclusion criteria for the final scoping review.

Severe pneumonia was defined according to the World Health Organization (WHO) guideline in 12 studies [3, 11–21]. The Italian study confirmed all patients as having severe pneumonia after contacting the author [22].

Eleven studies contained both severe and non-severe pneumonia [3, 11–20], while the remaining two studies exclusively included severe pneumonia [21, 22]. Most studies were retrospective, observational, and single-center-based [3, 11, 13–18, 20]. Most studies summarized both clinical characteristics and laboratory findings [3, 12–15, 17–21]. Two studies without available clinical data described information regarding features about cytokines and lymphocyte subsets [11, 16] (Table 1).

**Table 1.**
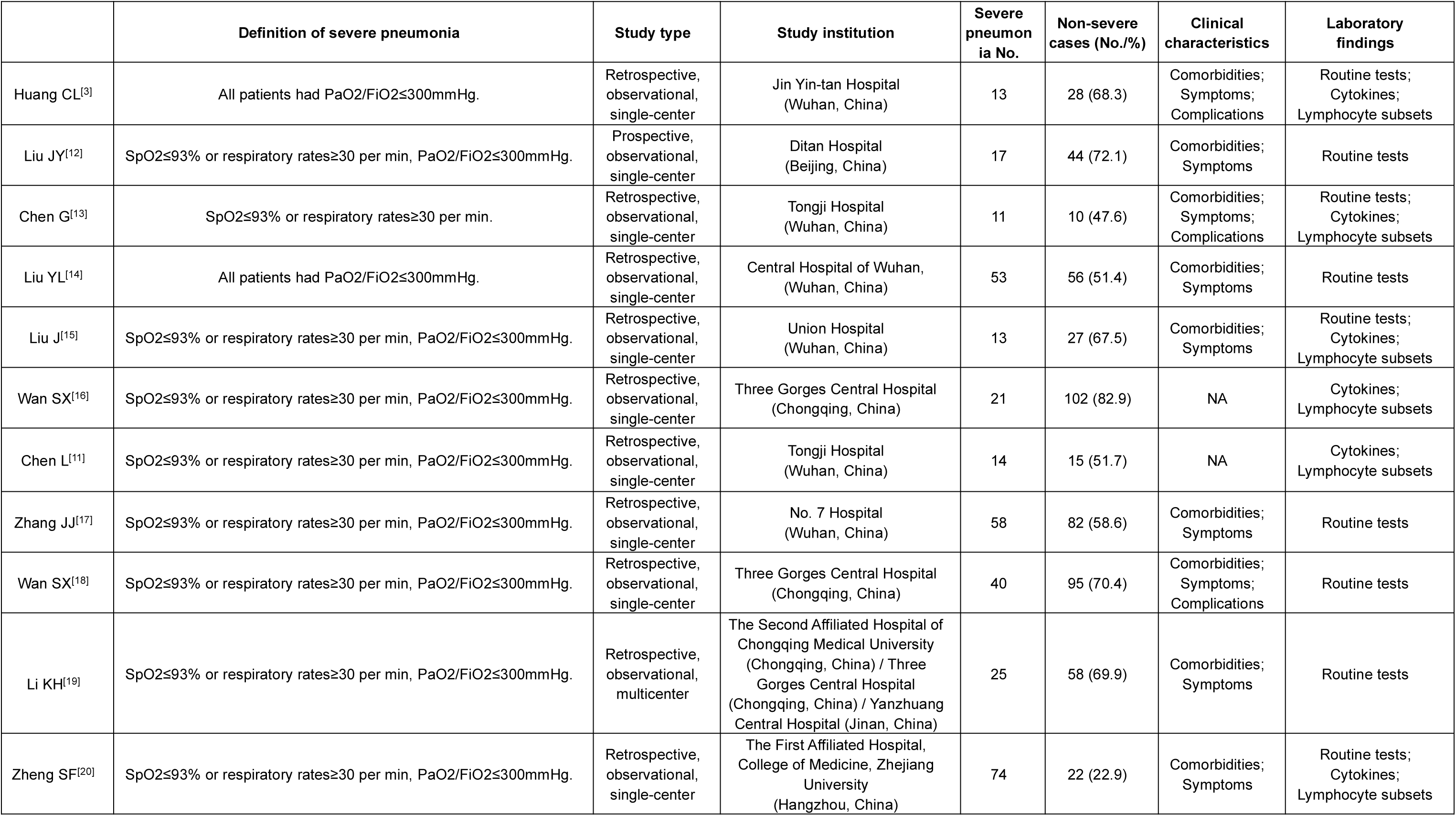
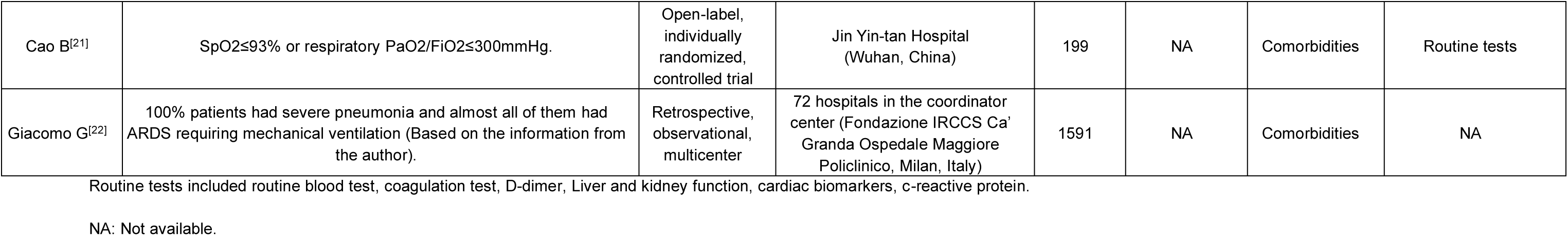
The characteristics of the included studies.

|  | Definition of severe pneumonia | Study type | Study institution | Severe pneumonia No. | Non-severe cases (No./%) | Clinical characteristics | Laboratory findings |
| --- | --- | --- | --- | --- | --- | --- | --- |
| Huang CL<sup>[3]</sup> | All patients had PaO<sub>2</sub>/FiO<sub>2</sub>≤300mmHg. | Retrospective, observational, single-center | Jin Yin-tan Hospital (Wuhan, China) | 13 | 28 (68.3) | Comorbidities; Symptoms; Complications | Routine tests; Cytokines; Lymphocyte subsets |
| Liu JY<sup>[12]</sup> | SpO<sub>2</sub>≤93% or respiratory rates≥30 per min, PaO<sub>2</sub>/FiO<sub>2</sub>≤300mmHg. | Prospective, observational, single-center | Ditan Hospital (Beijing, China) | 17 | 44 (72.1) | Comorbidities; Symptoms | Routine tests |
| Chen G<sup>[13]</sup> | SpO<sub>2</sub>≤93% or respiratory rates≥30 per min. | Retrospective, observational, single-center | Tongji Hospital (Wuhan, China) | 11 | 10 (47.6) | Comorbidities; Symptoms; Complications | Routine tests; Cytokines; Lymphocyte subsets |
| Liu YL<sup>[14]</sup> | All patients had PaO<sub>2</sub>/FiO<sub>2</sub>≤300mmHg. | Retrospective, observational, single-center | Central Hospital of Wuhan, (Wuhan, China) | 53 | 56 (51.4) | Comorbidities; Symptoms | Routine tests |
| Liu J<sup>[15]</sup> | SpO<sub>2</sub>≤93% or respiratory rates≥30 per min, PaO<sub>2</sub>/FiO<sub>2</sub>≤300mmHg. | Retrospective, observational, single-center | Union Hospital (Wuhan, China) | 13 | 27 (67.5) | Comorbidities; Symptoms | Routine tests; Cytokines; Lymphocyte subsets |
| Wan SX<sup>[16]</sup> | SpO<sub>2</sub>≤93% or respiratory rates≥30 per min, PaO<sub>2</sub>/FiO<sub>2</sub>≤300mmHg. | Retrospective, observational, single-center | Three Gorges Central Hospital (Chongqing, China) | 21 | 102 (82.9) | NA | Cytokines; Lymphocyte subsets |
| Chen L<sup>[11]</sup> | SpO<sub>2</sub>≤93% or respiratory rates≥30 per min, PaO<sub>2</sub>/FiO<sub>2</sub>≤300mmHg. | Retrospective, observational, single-center | Tongji Hospital (Wuhan, China) | 14 | 15 (51.7) | NA | Cytokines; Lymphocyte subsets |
| Zhang JJ<sup>[17]</sup> | SpO<sub>2</sub>≤93% or respiratory rates≥30 per min, PaO<sub>2</sub>/FiO<sub>2</sub>≤300mmHg. | Retrospective, observational, single-center | No. 7 Hospital (Wuhan, China) | 58 | 82 (58.6) | Comorbidities; Symptoms | Routine tests |
| Wan SX<sup>[18]</sup> | SpO<sub>2</sub>≤93% or respiratory rates≥30 per min, PaO<sub>2</sub>/FiO<sub>2</sub>≤300mmHg. | Retrospective, observational, single-center | Three Gorges Central Hospital (Chongqing, China) | 40 | 95 (70.4) | Comorbidities; Symptoms; Complications | Routine tests |
| Li KH<sup>[19]</sup> | SpO<sub>2</sub>≤93% or respiratory rates≥30 per min, PaO<sub>2</sub>/FiO<sub>2</sub>≤300mmHg. | Retrospective, observational, multicenter | The Second Affiliated Hospital of Chongqing Medical University (Chongqing, China) / Three Gorges Central Hospital (Chongqing, China) / Yanzhuang Central Hospital (Jinan, China) | 25 | 58 (69.9) | Comorbidities; Symptoms | Routine tests |
| Zheng SF<sup>[20]</sup> | SpO<sub>2</sub>≤93% or respiratory rates≥30 per min, PaO<sub>2</sub>/FiO<sub>2</sub>≤300mmHg. | Retrospective, observational, single-center | The First Affiliated Hospital, College of Medicine, Zhejiang University (Hangzhou, China) | 74 | 22 (22.9) | Comorbidities; Symptoms | Routine tests; Cytokines; Lymphocyte subsets |
| Cao B<sup>[21]</sup> | SpO<sub>2</sub>≤93% or respiratory PaO<sub>2</sub>/FiO<sub>2</sub>≤300mmHg. | Open-label, individually randomized, controlled trial | Jin Yin-tan Hospital (Wuhan, China) | 199 | NA | Comorbidities | Routine tests |
| Giacomo G<sup>[22]</sup> | 100% patients had severe pneumonia and almost all of them had ARDS requiring mechanical ventilation (Based on the information from the author). | Retrospective, observational, multicenter | 72 hospitals in the coordinator center (Fondazione IRCCS Ca' Granda Ospedale Maggiore Policlinico, Milan, Italy) | 1591 | NA | Comorbidities | NA |

Routine tests included routine blood test, coagulation test, D-dimer, Liver and kidney function, cardiac biomarkers, c-reactive protein.
NA: Not available.

### Sources of evidence appraisal

The quality assessment of individual sources of evidence by JBI tool was summarized for both case-series and cross-sectional studies. All studies had good quality and were processed for data summarization (Table S1-2).

### Demographics

All studies used in this review included demographics of patients with severe pneumonia [3, 11–22]. The percentages of male severe pneumonia patients ranged from 52.4% to 90.9% but no study claimed that male patients were at greater risk of developing SARS-CoV-2 pneumonia. The mean age of patients in each study ranged from 49 to 64 years old.

### Comorbidities

Eleven studies selected for review described comorbidities of severe pneumonia [3, 12–15, 17–22]. The most frequent comorbidity was hypertension (45.0% (8.0-48.8% by 10 studies) [3, 12–15, 17–20, 22], followed by diabetes (16.7% (7.7-30.8%) by 11 studies) [3, 12–15, 17–22], cardiovascular disease (17.1% (4.0-23.1%) by nine studies) [3, 12, 14, 17–22], and chronic pulmonary disease (4.5% (2.5-17.6%) by eight studies) [3, 12, 14, 17–20, 22]. Chronic kidney disease (3.8% (1.3-15.1%) by four studies) [14, 17, 20, 22], cerebrovascular disease (11.3% by one study) [14], and cancer (6.5% (0-7.8%) by six study) [3, 15, 18, 20–22] were also found in the patients with severe pneumonia (Table S3).

### Clinical manifestations

Nine of the selected studies described symptoms present in patients with severe pneumonia [3, 12–15, 17–20]. On admission, the most common symptom was fever (90.6% (79.2-100%) by nine studies) [3, 12–15, 17–20]. Additional common signs included cough (73.8% (56.8-96.0%) by nine studies) [3, 12–15, 17–20], fatigue (50.9% (47.5-100%) by eight studies) [3, 12–15, 17, 18, 20] and dyspnea (34.6% (0-100%) by eight studies) [3, 12, 13, 15, 17–20], only one study reported no dyspnea at admission in severe pneumonia [15]. Sputum production, headache and digestive symptoms were also reported. Less commonly recorded symptoms were myalgia, pharyngalgia, chest tightness, dizziness and anorexia (Table S4). As compared to non-severe pneumonia, severe pneumonia were more frequently featured with dyspnea [3, 12, 13, 19], cough [17, 19], sputum production [12, 15, 19], fatigue [14, 18], and nausea [17]. No difference of fever prevalence was found at admission.

### Complications

Four studies described complications regarding severe pneumonia [3, 13, 14, 18]. ARDS (79.5% (50.0-100%) by four studies) was the most common complication in severe pneumonia [3, 13, 14, 18]. Other fatal complications included acute cardiac injury (10.9% (5.0-30.8%) by three studies) [3, 13, 18], acute kidney injury (7.8% (2.5-23.1%) by three studies) [3, 13, 18] and shock (7.5% (2.5-23.1%) by two studies) [3, 18]. Additionally, liver dysfunction and disseminated intravascular coagulation were also reported [13] (Table S5).

### Laboratory findings

Of the 10 studies describing laboratory findings for severe pneumonia [3, 12–15, 17–21], nine of them including a comparison between severe and non-severe pneumonia [3, 12–15, 17–20]. We synthesized medians for white blood cell (WBC) count (4.5-11.3×10^9^/L by 10 studies) [3, 12–15, 17–21], neutrophil count (2.8-10.6×10^9^/L by seven studies) [3, 12–15, 18, 19], lymphocyte count (0.4-0.9×10^9^/L by 10 studies) [3, 12–15, 17–21], and lactate dehydrogenase (LDH) (264.0-567.2 U/L by seven studies) [3, 13–15, 18, 20, 21]. When compared with non-severe pneumonia, severe pneumonia had the lower lymphocyte count [3, 12–15, 17–20], higher levels of D-dimer [3, 13–15, 17, 18, 20], LDH [3, 13–15, 18, 20] and hypersensitive troponin I [3, 20] in all associated studies. Severe pneumonia presented with the higher levels of WBC [3, 13–15, 17], neutrophil [3, 12–15, 18], ALT [3, 13, 15], AST [13, 15, 18] and C-reactive protein [13-15, 17-, and lower albumin levels [3, 13, 18, 20] in the majority of related studies. The levels of platelet count, hemoglobin, prothrombin time, activated partial thromboplastin time, creatine kinase and creatine did not significantly differ between severe and non-severe pneumonia (Table S6).

Six studies reported levels of cytokines [3, 11, 13, 15, 16, 20], and four studies reported counts of lymphocyte subsets for severe pneumonia [13, 15, 16, 20]. We synthesized medians for IL-6 (7.1-73.8 pg/mL) [3, 11, 13, 15, 16, 20], IL-10 (4.6-11.1 pg/mL) [3, 11, 13, 15, 16, 20], CD8+ T cell (124.3-179.0×10^6^/L) [13, 15, 18, 20] and CD4+ T cell (171.5-275.7×10^6^/L) [13, 15, 18, 20]. As compared to non-severe pneumonia, severe pneumonia in most studies had higher levels of IL-6 [11, 15, 18] and IL-10 [3, 13, 15, 18], and lower CD8+ [13, 15, 18, 20] and CD4+ T [13, 18, 20] cell counts. The medians of TNF-alpha, B and NK cell count were not significantly different between severe and non-severe pneumonia (Table S7).

### Treatment

Eight studies described treatment for severe SARS-CoV-2 pneumonia [3, 12–14, 18, 20–22]. Antiviral therapy was commonly used for severe pneumonia (48.2-100% by six studies) [3, 12, 14, 18, 20, 21]. The most frequently used antiviral agent was oseltamivir (75 mg twice daily, orally), other antivirals used included lopinavir, ritonavir, remdesivir, favipiravir, and darunavir-cobicistat. Corticosteroid was widely used in China (33.7-93.2% by six studies) [3, 13, 14, 18, 20, 21], but was not reported as used in Italian study [22]. Other used treatments included immunoglobulin (54.7-66.2%) [14, 20], antibiotic agents (43.2-100%) [3, 12–14, 18, 20, 21], renal replacement therapy (4.5-23.1%) [3, 18, and oxygen support (nasal cannula (0.8-29.4%) [3, 12, 22], non-invasive ventilation or high flow nasal cannula (8.6-81.8%) [3, 12–14, 18, 21, 22], invasive mechanial ventilation (2.5-72.3%) [3, 12, 18, 20–22] and extracorporeal membrane oxygenation (0-15.3%) [3, 18, 20–22]). Ventilation therapy, especially mechanical ventilation, was the primary treatment for ARDS. The Italian study presented details reporting the positive end-expiratory pressure (PEEP) median was 14 cm H_2_O, and PEEP levels as high as 22 cm H_2_O were applied. When compared with younger patients (≤63 years), older patients had higher FiO_2_ and lower median PaO_2_/FIO_2_. No difference of PEEP was reported [22].

The therapeutic efficacy was evaluated or reported in three studies [14, 21, 23], Cao et al. reported no benefit was observed with lopinavir-ritonavir treatment beyond standard care [21], no benefit was found for survival probability when using ribavirin, oseltamivir, arbidol, corticosteroid, or immunoglobulin [14, 23] (Table 2).

**Table 2.**
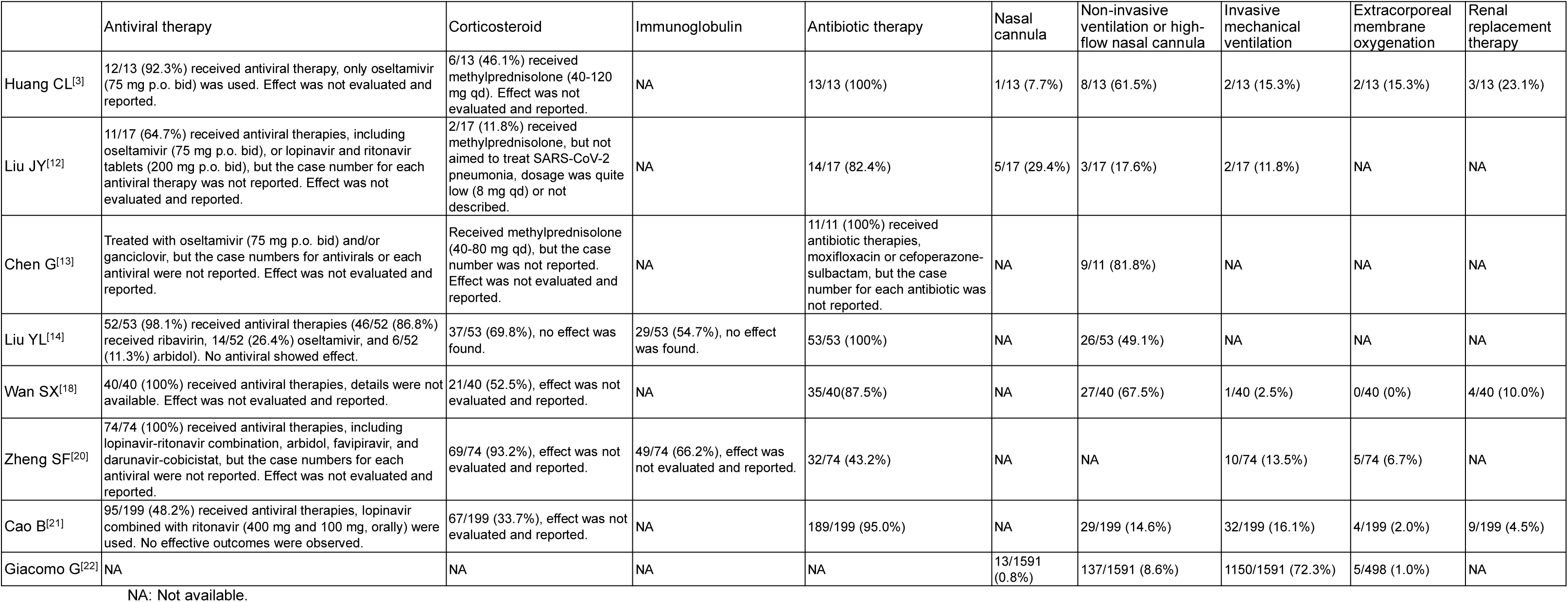
The treatment for severe SARS-CoV-2 pneumonia patients.

### Prognosis

The case-fatality rates for severe pneumonia varied from 15.4% to 49.1% in six studies with one to four weeks follow-up [3, 13–15, 21, 22].

## Discussion

This scoping review, the first systematic summarization of aspects from 2,129 patients of severe pneumonia in 13 studies, provided urgently needed information to frontline clinicians and allied health professionals involved in the care of severe pneumonia, and identified the knowledge gaps between what already known and unknown.

As for the management of severe pneumonia, there have been clinical guidelines released by WHO [8] and Surviving Sepsis Campaign (SSC) [24, 25]. However, the guidelines do not give any recommendation on the predisposing factors of disease severity or mortality in patients with severe pneumonia. In published literatures, studies on COVID-19 suggested old age and comorbidities as the predisposing factors of mortality [26, 27], but not exclusively observed in severe pneumonia. A retrospective cohort study, with the confounding adequately adjusted, found older age as a predictor of poor prognosis in 191 adult patients with COVID-19 [26]. Another study based on 150 patients of COVID-19, with the limitation such as unadjusted confounding in methodology, indicated cardiovascular disease as the clinical predictor of mortality [27]. In this scoping review, the older age and higher frequency of comorbidities such as chronic pulmonary disease, hypertension or diabetes in severe pneumonia compared to non-severe pneumonia might serve as the potential predisposing factors of disease severity, but reported with unadjusted estimates [14, 15, 17–20]. So, multicenter- and large population-designed studies, with careful delineation of disease severity (consistent definition, enough monitoring time), survivor or non-survivor categorization, and confounding adequately controlled, are required to define the predisposing factors that contribute to disease severity and mortality in severe pneumonia.

No recommendation on using laboratory examinations to predict disease severity or mortality in patients with severe pneumonia was given based on the guidelines. Current evidence from studies of patients with COVID-19 revealed lymphopenia as the predisposing factor of disease mortality [27, 28], and considered D-dimer as the risk factor of complications such as thrombosis [29] and septic shock [4, 26, 27]. In this review, the summarization of laboratory tests also might have some suggestions on the predisposing factors of disease severity. For example, the consistent existing of lymphopenia and increase of D-dimer and LDH in severe pneumonia across all studies suggests lymphopenia, D-dimer and LDH as the potential critical factors of disease severity, but needs validation from confounding-controlled study. The higher levels of IL-6 and IL-10 whereas lower counts of CD8+ and CD4+ T cells, due to the cytokine storm and exhausted immunity in severe pneumonia, also might be used for disease severity prediction [3, 30]. No evidence on utilizing laboratory tests for mortality or complication prediction in severe pneumonia according to this scoping review.

Recommendations regarding therapies for severe pneumonia have been listed by the guidelines [8, 24, 25]. Guidelines suggest against the routine use of antiviral agents in severe pneumonia treatment, as no study reported antivirals had a direct inhibitory effect in SARS-CoV-2 virus replication [31, 32] or provided clinical benefit in COVID-19 patients in randomized controlled trials (RCT) [21, 33]. An open-label RCT with 199 patients did not observe a benefit from lopinavir-ritonavir treatment beyond standard care [21]. Another recent multicenter-based RCT found remdesivir was not associated with statistically significant clinical benefits in severe pneumonia [33]. Our scoping review also does not support the routine use of antivirals due to the lack of observed efficacy from empirically using antivirals at the beginning of pandemic. Therefore, we are expecting other ongoing RCT to confirm or exclude a benefit from antivirals for severe pneumonia. The routine use of corticosteroid is not recommended in COVID-19 treatment by the guidelines [8, 24, 25], due to the lack of effectiveness and possible harm such as delayed viral clearance based on the experience of using corticosteroids for SARS [1] and MERS treatment [34, 35]. However, Chinese clinicians thought the unrecommended use of corticosteroid was mostly based on the findings from observational and inconclusive studies, which they thought should not be a reason for abandoning corticosteroid use in COVID-19 [36, 37]. A recent study result from University of Oxford, but have not been published or subjected to scientific scrutiny, showed low dose of dexamethasone (6 mg for 10 days) significantly reduced deaths of severely sick COVID-19 patients in a major clinical trial [38]. In our scoping review, a higher dose of corticosteroid has been widely used in severe pneumonia in selected Chinese studies, but needs RCT to gain more solid and conclusive evidence. Other treatments for severe pneumonia suggested by our review and the guidelines contain empiric antimicrobial treatment [39–42] and mechanical ventilation, nevertheless, details such as opportunity for advanced oxygen/ventilatory support, tidal volumes and inspiratory pressure, and prone ventilation were not presented in most selected studies.

Other findings in this review might have some new implications on therapeutic strategy for severe pneumonia. The cytokine storm identified in severe pneumonia might provide a reason and basis for continuous renal replacement therapy or corticosteroids administration, but the effect on outcomes needs to be validated. The fluid resuscitation strategy for patients with severe pneumonia should be carefully modified according to our findings regarding the complications, as in the setting of severe pneumonia-induced ARDS, acute cardiac injury, and acute kidney injury, fluid resuscitation might increase the risks of pulmonary edema, heart failure and kidney failure if using starches, respectively.

Several limitations need to be acknowledged. First, some features of severe pneumonia were noted in small numbers of studies, and the majority of studies included small numbers of patients, so, the features such as cerebrovascular disease, pharyngalgia, and hypersensitive troponin I might be unrepresentative, and possibly lead to uncertainty in practical guidance. Second, the inherent risk of confounding in the included studies would be high, as the majority of studies were observational. Lastly, there was a wide range of presentations recorded in the studies overall. To have a comprehensive understanding on the features of severe pneumonia, we delineated the aspects with the minimum and maximum of the categorical data, and calculated overall ratio of each item in comorbidities, symptoms and complications. In future, there is a need for systematic inclusion of many points of data.

Collectively, this scoping review gives us a profiling regarding the demographics, comorbidities, clinical manifestations, complications, laboratory findings, therapies, and outcomes of severe pneumonia. The old age, some comorbidities and laboratory findings might be the predisposing factors of disease severity. Most Chinese studies were performed at the beginning of pandemic and ahead of the clear acknowledgement of COVID-19, the empirical therapy such as antivirals were used as the primary treatment for severe pneumonia. Also, as most data originate from China, the external validity of findings need to be confirmed with the studies from western countries. So, to better understand the disease and provide the guidance to treatment, multicenter- and large population-designed studies, with confounding variables controlled and long enough to accommodate follow-ups are urgently required in the future.

## Data Availability

All data produced in the present study are available upon reasonable request to the authors

## Declarations

### Authors’ contributions

J. Huang was the guarantor of the entire manuscript for designing and supervising the entire study. Analysis and interpretation of the data: X. Li, H. Zhang, J. Huang. Drafting of the article: X. Li, Z. Cai. Critical revision of the article for important intellectual content: X. Li, H. Zhang, J. Huang, and G. Ren. Final approval of the article: X. Li, H. Zhang, Z. Cai, G. Ren, J. Huang. Collection and assembly of data: X. Li, H. Zhang, Z. Cai.

---

**Table S1.**
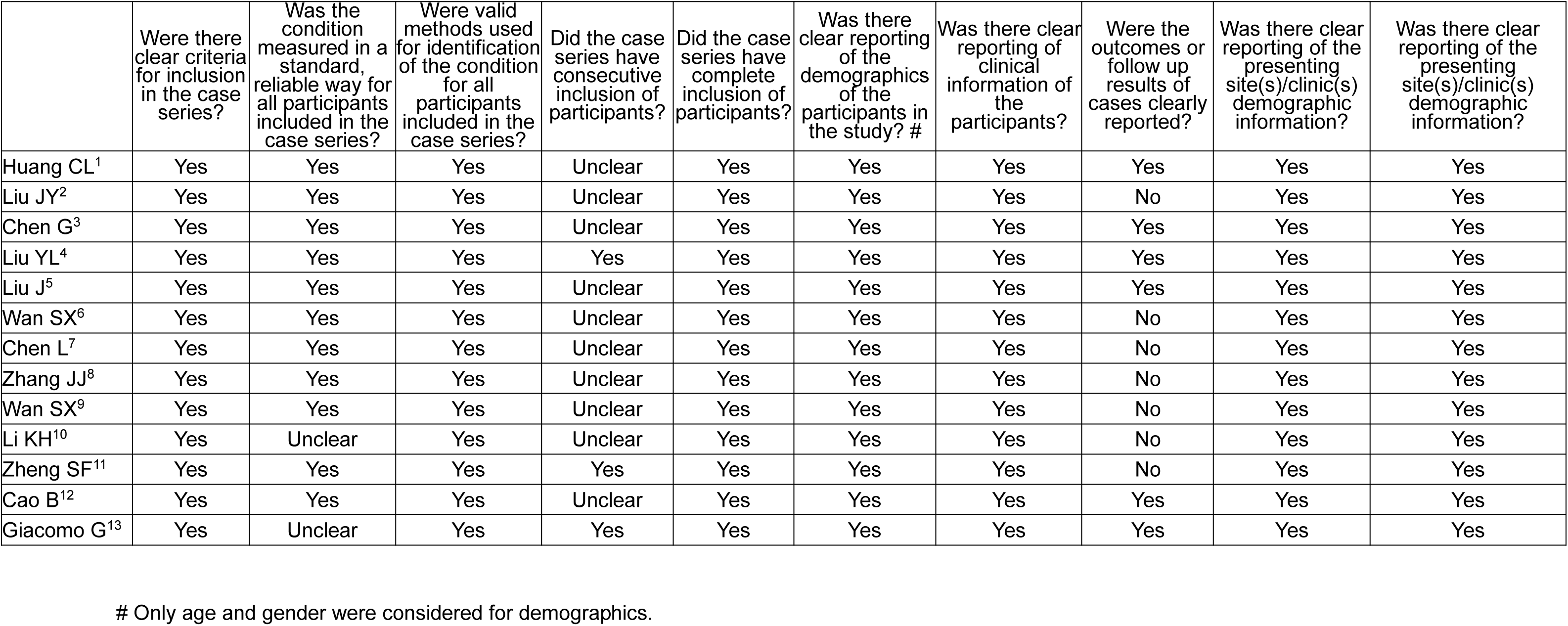
Critical appraisal for case-series.

**Table S2.** Critical appraisal for cross-sectional studies.

|  | Were the criteria for inclusion in the sample clearly defined? | Were the study subjects and the setting described in detail? | Was the exposure measured in a valid and reliable way? | Were objective, standard criteria used for measurement of the condition? | Were confounding factors identified? | Were strategies to deal with confounding factors stated? | Were the outcomes measured in a valid and reliable way? | Was appropriate statistical analysis used? |
| --- | --- | --- | --- | --- | --- | --- | --- | --- |
| Huang CL<sup>1</sup> | Yes | Yes | Yes | Yes | No | No | Yes | Yes |
| Liu JY<sup>2</sup> | Yes | Yes | Yes | Yes | No | No | No | Yes |
| Chen G<sup>3</sup> | Yes | Yes | Yes | Yes | No | No | Yes | Yes |
| Liu YL<sup>4</sup> | Yes | Yes | Yes | Yes | No | No | Yes | Yes |
| Liu J<sup>5</sup> | Yes | Yes | Yes | Yes | No | No | Yes | Yes |
| Wan SX<sup>6</sup> | Yes | Yes | Yes | Yes | No | No | No | Yes |
| Chen L<sup>7</sup> | Yes | Yes | Yes | Yes | No | No | No | Yes |
| Zhang JJ<sup>8</sup> | Yes | Yes | Yes | Yes | No | No | No | Yes |
| Wan SX<sup>9</sup> | Yes | Yes | Yes | Yes | No | No | No | Yes |
| Li KH<sup>10</sup> | Yes | Yes | Yes | Yes | No | No | No | Yes |
| Zheng SF<sup>11</sup> | Yes | Yes | Yes | Yes | No | No | No | Yes |

**Table S3.** The comorbidities of severe SARS-CoV-2 pneumonia patients.

|  | Any comorbidity | Hypertension | Diabetes | Cardiovascular disease | Chronic pulmonary disease | Chronic kidney disease | Cerebrovascular disease | Cancer | Immunodeficiency |
| --- | --- | --- | --- | --- | --- | --- | --- | --- | --- |
| Huang CL<sup>1</sup> | 5/13 (38.5%) | 2/13 (15.4%) | 1/13 (7.7%) | 3/13 (23.1%) | 1/13 (7.7%) | NA | NA | 0/13 (0%) | NA |
| Liu JY<sup>2</sup> | NA | 6/17 (35.3%) | 3/17 (17.6%) | 1/17 (5.9%) | 3/17 (17.6%) | NA | NA | NA | NA |
| Chen G<sup>3</sup> | 5/11 (45.5%) | 4/11 (36.4%) | 2/11 (18.2%) | NA | NA | NA | NA | NA | NA |
| Liu YL<sup>4</sup> | NA | 21/53 (39.6%) | 11/53 (20.8%)<sup>#</sup> | 3/53 (5.7%) | 2/53 (3.8%) | 8/53 (15.1%)<sup>#</sup> | 6/53 (11.3%)<sup>#</sup> | NA | NA |
| Liu J<sup>5</sup> | 7/13 (53.8%) | 5/13 (38.5%)<sup>#</sup> | 4/13 (30.8%) | NA | NA | NA | NA | 0/13 (0%) | NA |
| Zhang JJ<sup>8</sup> | 46/58 (79.3%)<sup>#</sup> | 22/58 (37.9%) | 8/58 (13.8%) | 4/58 (6.9%) | 2/58 (3.4%) | 2/58 (3.4%) | NA | NA | NA |
| Wan SX<sup>9</sup> | 28/40 (70.0%)<sup>#</sup> | 4/40 (10.0%) | 9/40 (22.5%) | 6/40 (15.0%) | 1/40 (2.5%) | NA | NA | 3/40 (7.5%) | NA |
| Li KH<sup>10</sup> | 11/25 (44.0%)<sup>#</sup> | 2/25 (8.0%) | 7/25 (28.0%)<sup>#</sup> | 1/25 (4.0%) | 4/25 (16.0%)<sup>#</sup> | NA | NA | NA | NA |
| Zheng SF<sup>11</sup> | NA | 31/74 (41.9%)<sup>#</sup> | 10/74 (13.5%) | 7/74 (9.4%) | 4/74 (5.4%) | 1/74 (1.3%) | NA | 1/74 (1.3%) | 1/74 (1.3%) |
| Cao B<sup>12</sup> | NA | NA | 23/199 (11.6%) | 13/199 (6.5%) | NA | NA | NA | 6/199 (3.0%) | NA |
| Giacomo G<sup>13</sup> | 709/1043 (68.0%) | 509/1043 (48.8%) | 180/1043 (17.3%) | 223/1043 (21.4%) | 42/1043 (4.0%) | 36/1043 (3.4%) | NA | 81/1043 (7.8%) | NA |
| **Overall ratio (Min-Max)** | 67.4%<br>(38.5-79.3%) | 45.0%<br>(8.0-48.8%) | 16.7%<br>(7.7-30.8%) | 17.1%<br>(4.0-23.1%) | 4.5%<br>(2.5-17.6%) | 3.8%<br>(1.3-15.1%) | 11.3% | 6.5%<br>(0-7.8%) | 1.3% |

NA: Not available; Min: Minimum; Max: Maximum; #: The comorbidity ratio was significantly different between severe and non-severe pneumonia

**Table S4.**
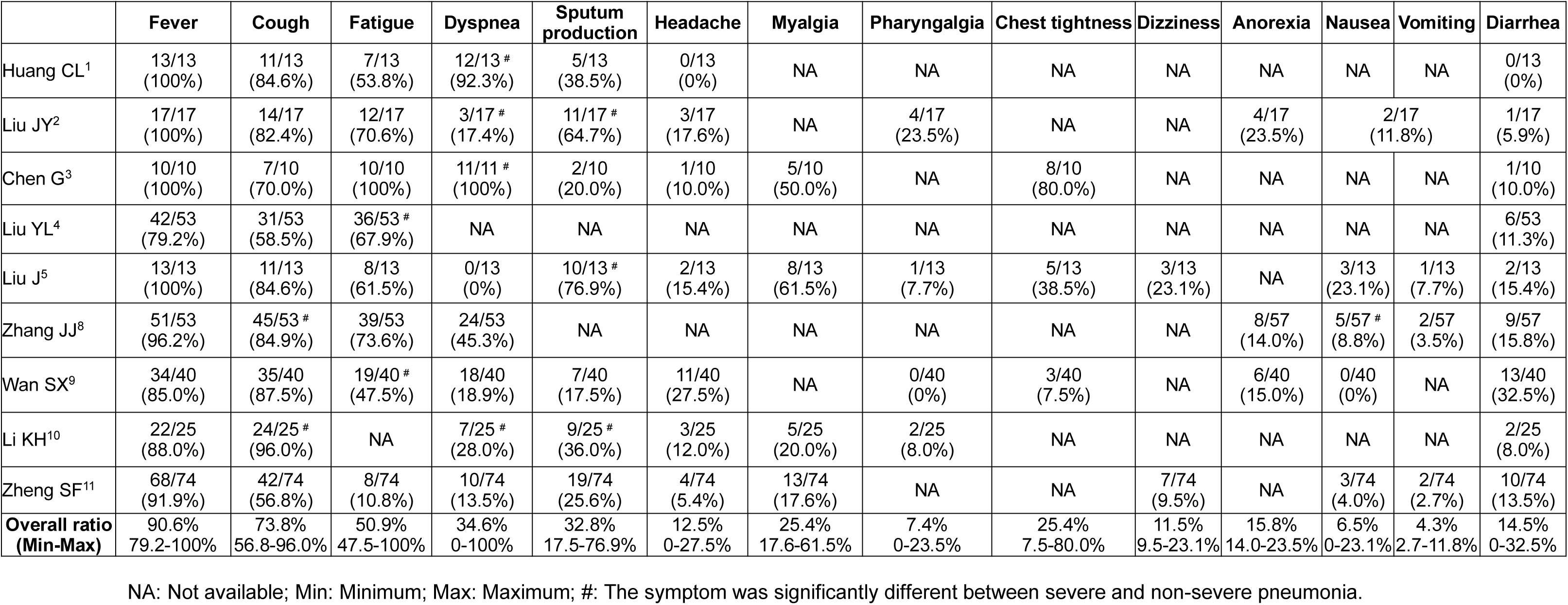
The symptoms of severe SARS-CoV-2 pneumonia patients.

**Table S5.** The complications of severe SARS-CoV-2 pneumonia patients.

|  | **Acute respiratory distress syndrome** | **Acute cardiac injury** | **Acute kidney injury** | **Shock** | **Liver dysfunction** | **Disseminated intravascular coagulation** |
| --- | --- | --- | --- | --- | --- | --- |
| Huang CL<sup>1</sup> | 11/13 (84.6%) # | 4/13 (30.8%) # | 3/13 (23.1%) # | 3/13 (23.1%) # | NA | NA |
| Chen G<sup>3</sup> | 9/11 (81.8%) # | 1/11 (9.1%) | 1/11 (9.1%) | NA | 2/11 (18.2%) | 1/11 (9.1%) |
| Liu YL<sup>4</sup> | 53/53 (100%) | NA | NA | NA | NA | NA |
| Wan SX<sup>9</sup> | 20/40 (50.0%) # | 2/40 (5.0%) | 1/40 (2.5%) | 1/40 (2.5%) | NA | NA |
| **Overall ratio (Min-Max)** | 79.5%<br>50.0-100% | 10.9%<br>5.0-30.8% | 7.8%<br>2.5-23.1% | 7.5%<br>2.5-23.1% | 18.2% | 9.1% |

NA: Not available; #: The prevalence of complications was significantly different between severe and non-severe pneumonia.

**Table S6.** The laboratory findings of severe SARS-CoV-2 pneumonia patients.

|  | Huang CL<sup>1</sup> | Liu JY<sup>2</sup> | Chen G<sup>3</sup> | Liu YL<sup>4</sup> | Liu J<sup>5</sup> | Zhang JJ<sup>8</sup> | Wan SX<sup>9</sup> | Li KH<sup>10</sup> | Zheng SF<sup>11</sup> | Cao B<sup>12</sup> |
| --- | --- | --- | --- | --- | --- | --- | --- | --- | --- | --- |
| White blood cell count, 10<sup>9</sup>/L | 11.3 (5.8-12.1) <sup>#</sup> | 4.5 (3.7-5.0) | 9.2±3.6 <sup>#</sup> | 5.4 (4.5-8.0) | 6.6 ± 3.4 <sup>#</sup> | 5.3 (4.0-9.0) <sup>#</sup> | 5.2 (4.9-6.9) | 5.5 (4.1-7.7) | 6.2 (3.9-10.8) | 7.0 (5.1–9.4) |
| Neutrophil count, x10<sup>9</sup>/L | 10.6 (5.0-11.8) <sup>#</sup> | 2.8 (2.3-4.4) <sup>#</sup> | 8.0±3.7 <sup>#</sup> | 4.1 (3.1-7.1) <sup>#</sup> | 4.7 (3.6-5.8) <sup>#</sup> | NA | 4.1 (3.1-5.7) <sup>#</sup> | 4.4 (2.9-6.5) | NA | NA |
| Lymphocyte count, x10<sup>9</sup>/L | 0.4 (0.2-0.8) <sup>#</sup> | 0.9 (0.7-1.1) <sup>#</sup> | 0.7±0.3 <sup>#</sup> | 0.7 (0.4-1.1) <sup>#</sup> | 0.6 (0.6-0.8) <sup>#</sup> | 0.7 (0.5-1.0) <sup>#</sup> | 0.8 (0.6-1.0) <sup>#</sup> | 0.7 (0.4-0.9) <sup>#</sup> | 0.7 (0.5-1.1) <sup>#</sup> | 0.9 (0.6–1.2) |
| Platelet count, x10<sup>9</sup>/L | 196.0 (165.0-263.0) | 153.0 (120.5-216.0) | 164.2±45.8 | NA | 186.6 ± 68.1 | NA | 147.0 (118.0-213.0) <sup>#</sup> | NA | 181 (140-227) | 207.0 (158.0–284.0) |
| Hemoglobin, g/L | 122.0 (111.0-128.0) | 138.0 (127.5-148.0) | 138.1±15.9 | NA | 123.4 ± 14.0 | NA | 130.0 (120.0-143.0) | NA | 135 (121-150) | NA |
| Prothrombin time, s | 12.2 (11.2-13.4) <sup>#</sup> | 12.0 (11.7-12.6) | 14.1±0.9 | NA | 13.4 ± 0.6 | NA | 11.3 (10.7-11.8) <sup>#</sup> | NA | NA | NA |
| Activated partial thromboplastin time, s | 26.2 (22.5-33.9) | NA | 36.2±5.8 <sup>#</sup> | NA | 39.5 ± 4.2 | NA | 29.7 (22.6-39.4) <sup>#</sup> | NA | NA | NA |
| D-dimer, mg/L | 2.4 (0.6-14.4) <sup>#</sup>,<br>Ref. 0-1.5 ug/L | NA | 8.2±9 ug/ml <sup>#</sup> | 940 (470-1905) <sup>#</sup>,<br>Ref. 0-500 mg/L | 0.9 (0.7-1.5) mg/L <sup>#</sup> | 0.4 (0.2-2.4) <sup>#</sup> | 0.6 (0.4-1.1) <sup>#</sup> | NA | 429 (260.5-787.5) ug/IFEU <sup>#</sup> | NA |
| Creatine kinase, U/L | 132.0 (82.0-493.0) | 119.0 (55.5-190.0) | 208.3±122.4 | 83 (49-169) | 104.0 (77.0-124.0) <sup>#</sup> | NA | 82.0 (56.3-146.2) <sup>#</sup> | NA | 78 (52-138) <sup>#</sup> | 69.0 (44.0-115.0) |
| Albumin, g/L | 27.9 (26.3-30.9) <sup>#</sup> | 43.0 (37.0-45.5) | 31.5±5.5 <sup>#</sup> | NA | NA | NA | 36.0 (33.0-38.5) <sup>#</sup> | NA | 37.9 (33.9-42.7) <sup>#</sup> | NA |
| Lactate dehydrogenase, U/L | 400.0 (323.0-578.0) <sup>#</sup> | NA | 567.2±217.1 <sup>#</sup> | 264 (190-448) <sup>#</sup> | 462.4 ± 190.6 <sup>#</sup> | NA | 309.0 (253.8-408.3) <sup>#</sup> | NA | 292.5 (227.3-355.8) <sup>#</sup> | 325.0 (245.0–433.0) |
| Alanine aminotransferase, U/L | 49.0 (29.0-115.0) <sup>#</sup> | 24.0 (14.0-34.5) | 41.4±14.9 <sup>#</sup> | 24 (16-31) | 27.0 (23.0-50.0) <sup>#</sup> | NA | 26.6 (14.5-33.3) | NA | 21 (14-27.5) | 33.0 (22.0–55.0) |
| Aspartate aminotransferase, U/L | 44.0 (30.0-70.0) | NA | 51±28.3 <sup>#</sup> | 31 (25-44) | 51.2 ± 18.7 <sup>#</sup> | NA | 33.6 (25.7-44.2) <sup>#</sup> | NA | 23 (18-34.5) | 34.0 (26.0–45.0) |
| Creatinine, umol/L | 79.0 (53.1-92.7) | 64.0 (51.0-81.0) | 90.7±38.5 | 67 (55-83) | 74.2 ± 28.3 | 66.0 (38.5-144.0) | 63.5 (51.5-74.3) | NA | 79 (65-91.5) <sup>#</sup> | 69.5 (57.2-82.5) |
| Hypersensitive troponin I, pg/mL | 3.3 (3.0-163.0) <sup>#</sup> | NA | NA | NA | NA | NA | NA | NA | 0.004 (0.002-0.01) ng/mL <sup>#</sup> | NA |
| C-reactive protein, mg/L | NA | 23.5 (2.7-71.2) | 135.2±52.4 <sup>#</sup><br>(hypersensitive) | 4.9 (2.7-6.5) <sup>#</sup> | 62.9 (42.4-86.6) <sup>#</sup> | 47.6 (20.6-87.1) <sup>#</sup> | 91.0 (52.7-136.3) <sup>#</sup> | 89.2 (47.9-134.6) <sup>#</sup> | 39.9 (16.2-63.7) <sup>#</sup> | NA |

NA: Not available; #: The laboratory finding was significantly changed between severe and non-severe pneumonia.

**Table S7.**
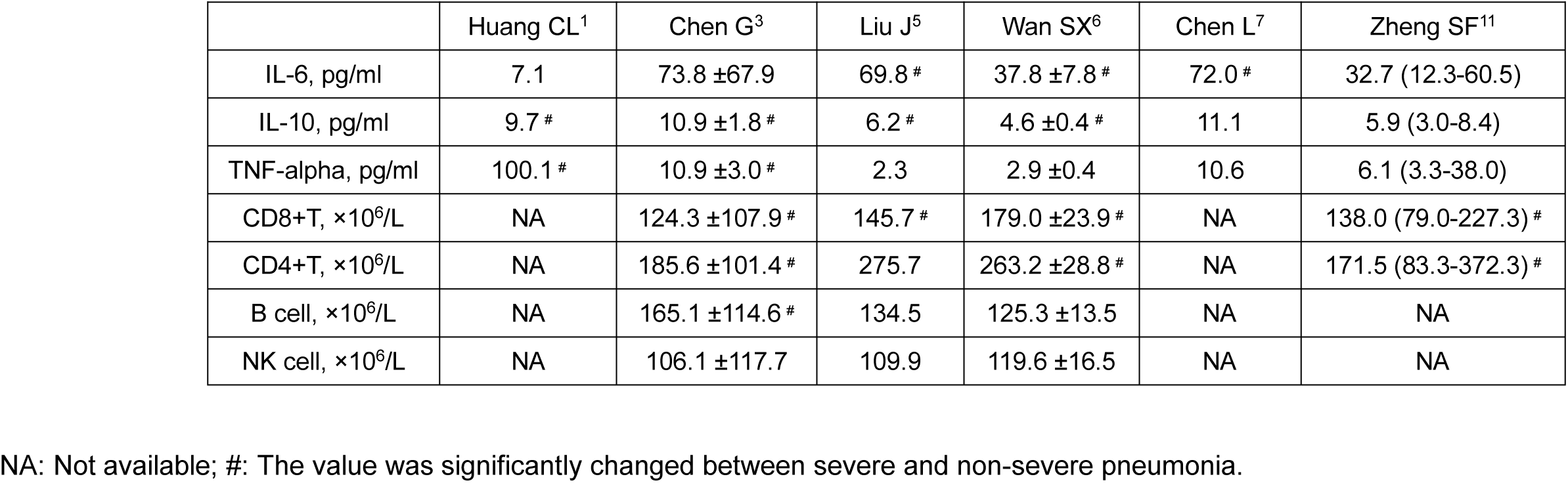
The cytokines and lymphocyte subsets of severe SARS-CoV-2 pneumonia patients.

|  | Huang CL<sup>1</sup> | Chen G<sup>3</sup> | Liu J<sup>5</sup> | Wan SX<sup>6</sup> | Chen L<sup>7</sup> | Zheng SF<sup>11</sup> |
| --- | --- | --- | --- | --- | --- | --- |
| IL-6, pg/ml | 7.1 | 73.8 ±67.9 | 69.8 # | 37.8 ±7.8 # | 72.0 # | 32.7 (12.3-60.5) |
| IL-10, pg/ml | 9.7 # | 10.9 ±1.8 # | 6.2 # | 4.6 ±0.4 # | 11.1 | 5.9 (3.0-8.4) |
| TNF-alpha, pg/ml | 100.1 # | 10.9 ±3.0 # | 2.3 | 2.9 ±0.4 | 10.6 | 6.1 (3.3-38.0) |
| CD8+T, ×10<sup>6</sup>/L | NA | 124.3 ±107.9 # | 145.7 # | 179.0 ±23.9 # | NA | 138.0 (79.0-227.3) # |
| CD4+T, ×10<sup>6</sup>/L | NA | 185.6 ±101.4 # | 275.7 | 263.2 ±28.8 # | NA | 171.5 (83.3-372.3) # |
| B cell, ×10<sup>6</sup>/L | NA | 165.1 ±114.6 # | 134.5 | 125.3 ±13.5 | NA | NA |
| NK cell, ×10<sup>6</sup>/L | NA | 106.1 ±117.7 | 109.9 | 119.6 ±16.5 | NA | NA |

NA: Not available; #: The value was significantly changed between severe and non-severe pneumonia.

## References

1. Stockman LJ, Bellamy R, Garner P, (2006) SARS: systematic review of treatment effects. PLoS Med 3: e343

2. 2. World Health Organization. COVID-19 situation report. Accessed June 4, 2020 https://covid19.who.int/

3. Huang C, Wang Y, Li X, Ren L, Zhao J, Hu Y, Zhang L, Fan G, Xu J, Gu X, Cheng Z, Yu T, Xia J, Wei Y, Wu W, Xie X, Yin W, Li H, Liu M, Xiao Y, Gao H, Guo L, Xie J, Wang G, Jiang R, Gao Z, Jin Q, Wang J, Cao B, (2020) Clinical features of patients infected with 2019 novel coronavirus in Wuhan, China. Lancet 395: 497–506

4. Yang X, Yu Y, Xu J, Shu H, Xia J, Liu H, Wu Y, Zhang L, Yu Z, Fang M, Yu T, Wang Y, Pan S, Zou X, Yuan S, Shang Y, (2020) Clinical course and outcomes of critically ill patients with SARS-CoV-2 pneumonia in Wuhan, China: a single-centered, retrospective, observational study. Lancet Respir Med 8: 475–481

5. Liao X, Wang B, Kang Y, (2020) Novel coronavirus infection during the 2019-2020 epidemic: preparing intensive care units-the experience in Sichuan Province, China. Intensive Care Med 46: 357–360

6. Arksey H, O’Malley L, (2005) Scoping studies: towards a methodological framework. International Journal of Social Research Methodology 8: 19–32

7. Tricco AC, Lillie E, Zarin W, O’Brien KK, Colquhoun H, Levac D, Moher D, Peters MDJ, Horsley T, Weeks L, Hempel S, Akl EA, Chang C, McGowan J, Stewart L, Hartling L, Aldcroft A, Wilson MG, Garritty C, Lewin S, Godfrey CM, Macdonald MT, Langlois EV, Soares-Weiser K, Moriarty J, Clifford T, Tuncalp O, Straus SE, (2018) PRISMA Extension for Scoping Reviews (PRISMA-ScR): Checklist and Explanation. Ann Intern Med 169: 467–473

8. WHO., Clinical management of severe acute respiratory infection (SARI) when COVID-19 disease is suspected. Mar 13, 2020 https://wwwwhoint/publications-detail/clinical-management-of-severe-acute-respiratory-infection-when-novel-coronavirus-(ncov)-infection-is-suspected (accessed Mar 22, 2020)

9. Force ADT, Ranieri VM, Rubenfeld GD, Thompson BT, Ferguson ND, Caldwell E, Fan E, Camporota L, Slutsky AS, (2012) Acute respiratory distress syndrome: the Berlin Definition. JAMA 307: 2526–2533

10. Jordan Z, Lockwood C, Munn Z, Aromataris E, (2019) The updated Joanna Briggs Institute Model of Evidence-Based Healthcare. Int J Evid Based Healthc 17: 58–71

11. Chen L, Liu HG, Liu W, Liu J, Liu K, Shang J, Deng Y, Wei S, (2020) [Analysis of clinical features of 29 patients with 2019 novel coronavirus pneumonia]. Chinese journal of tuberculosis and respiratory diseases 43: E005

12. Liu J, Liu Y, Xiang P, Pu L, Xiong H, Li C, Zhang M, Tan J, Xu Y, Song R, Song M, Wang L, Zhang W, Han B, Yang L, Wang X, Zhou G, Zhang T, Li B, Wang Y, Chen Z, Wang X, (2020) Neutrophil-to-Lymphocyte Ratio Predicts Severe Illness Patients with 2019 Novel Coronavirus in the Early Stage. medRxiv 10.1101/2020.02.10.20021584: 2020.2002.2010.20021584

13. Chen G, Wu D, Guo W, Cao Y, Huang D, Wang H, Wang T, Zhang X, Chen H, Yu H, Zhang X, Zhang M, Wu S, Song J, Chen T, Han M, Li S, Luo X, Zhao J, Ning Q, (2020) Clinical and immunological features of severe and moderate coronavirus disease 2019. The Journal of Clinical Investigation 130: 2620–2629

14. Liu Y, Sun W, Li J, Chen L, Wang Y, Zhang L, Yu L, (2020) Clinical features and progression of acute respiratory distress syndrome in coronavirus disease 2019. medRxiv 10.1101/2020.02.17.20024166: 2020.2002.2017.20024166

15. Liu J, Li S, Liu J, Liang B, Wang X, Wang H, Li W, Tong Q, Yi J, Zhao L, Xiong L, Guo C, Tian J, Luo J, Yao J, Pang R, Shen H, Peng C, Liu T, Zhang Q, Wu J, Xu L, Lu S, Wang B, Weng Z, Han C, Zhu H, Zhou R, Zhou H, Chen X, Ye P, Zhu B, Wang L, Zhou W, He S, He Y, Jie S, Wei P, Zhang J, Lu Y, Wang W, Zhang L, Li L, Zhou F, Wang J, Dittmer U, Lu M, Hu Y, Yang D, Zheng X, (2020) Longitudinal characteristics of lymphocyte responses and cytokine profiles in the peripheral blood of SARS-CoV-2 infected patients. EBioMedicine 10.1016/j.ebiom.2020.102763: 102763

16. Wan S, Yi Q, Fan S, Lv J, Zhang X, Guo L, Lang C, Xiao Q, Xiao K, Yi Z, Qiang M, Xiang J, Zhang B, Chen Y, (2020) Characteristics of lymphocyte subsets and cytokines in peripheral blood of 123 hospitalized patients with 2019 novel coronavirus pneumonia (NCP). medRxiv 10.1101/2020.02.10.20021832: 2020.2002.2010.20021832

17. Zhang JJ, Dong X, Cao YY, Yuan YD, Yang YB, Yan YQ, Akdis CA, Gao YD, (2020) Clinical characteristics of 140 patients infected with SARS-CoV-2 in Wuhan, China. Allergy 10.1111/all.14238

18. Wan S, Xiang Y, Fang W, Zheng Y, Li B, Hu Y, Lang C, Huang D, Sun Q, Xiong Y, Huang X, Lv J, Luo Y, Shen L, Yang H, Huang G, Yang R, (2020) Clinical features and treatment of COVID-19 patients in northeast Chongqing. J Med Virol 10.1002/jmv.25783

19. Li K, Wu J, Wu F, Guo D, Chen L, Fang Z, Li C, (2020) The Clinical and Chest CT Features Associated with Severe and Critical COVID-19 Pneumonia. Invest Radiol 10.1097/RLI.0000000000000672

20. Zheng S, Fan J, Yu F, Feng B, Lou B, Zou Q, Xie G, Lin S, Wang R, Yang X, Chen W, Wang Q, Zhang D, Liu Y, Gong R, Ma Z, Lu S, Xiao Y, Gu Y, Zhang J, Yao H, Xu K, Lu X, Wei G, Zhou J, Fang Q, Cai H, Qiu Y, Sheng J, Chen Y, Liang T, (2020) Viral load dynamics and disease severity in patients infected with SARS-CoV-2 in Zhejiang province, China, January-March 2020: retrospective cohort study. BMJ 369: m1443

21. 21. Cao B, Wang Y, Wen D, Liu W, Wang J, Fan G, Ruan L, Song B, Cai Y, Wei M, Li X, Xia J, Chen N, Xiang J, Yu T, Bai T, Xie X, Zhang L, Li C, Yuan Y, Chen H, Li H, Huang H, Tu S, Gong F, Liu Y, Wei Y, Dong C, Zhou F, Gu X, Xu J, Liu Z, Zhang Y, Li H, Shang L, Wang K, Li K, Zhou X, Dong X, Qu Z, Lu S, Hu X, Ruan S, Luo S, Wu J, Peng L, Cheng F, Pan L, Zou J, Jia C, Wang J, Liu X, Wang S, Wu X, Ge Q, He J, Zhan H, Qiu F, Guo L, Huang C, Jaki T, Hayden FG, Horby PW, Zhang D, Wang C, (2020) A Trial of Lopinavir-Ritonavir in Adults Hospitalized with Severe Covid-19. N Engl J Med 10.1056/NEJMoa2001282

22. Grasselli G, Zangrillo A, Zanella A, Antonelli M, Cabrini L, Castelli A, Cereda D, Coluccello A, Foti G, Fumagalli R, Iotti G, Latronico N, Lorini L, Merler S, Natalini G, Piatti A, Ranieri MV, Scandroglio AM, Storti E, Cecconi M, Pesenti A, Network C-LI, Nailescu A, Corona A, Zangrillo A, Protti A, Albertin A, Forastieri Molinari A, Lombardo A, Pezzi A, Benini A, Scandroglio AM, Malara A, Castelli A, Coluccello A, Micucci A, Pesenti A, Sala A, Alborghetti A, Antonini B, Capra C, Troiano C, Roscitano C, Radrizzani D, Chiumello D, Coppini D, Guzzon D, Costantini E, Malpetti E, Zoia E, Catena E, Agosteo E, Barbara E, Beretta E, Boselli E, Storti E, Harizay F, Della Mura F, Lorini FL, Donato Sigurta F, Marino F, Mojoli F, Rasulo F, Grasselli G, Casella G, De Filippi G, Castelli G, Aldegheri G, Gallioli G, Lotti G, Albano G, Landoni G, Marino G, Vitale G, Battista Perego G, Evasi G, Citerio G, Foti G, Natalini G, Merli G, Sforzini I, Bianciardi L, Carnevale L, Grazioli L, Cabrini L, Guatteri L, Salvi L, Dei Poli M, Galletti M, Gemma M, Ranucci M, Riccio M, Borelli M, Zambon M, Subert M, Cecconi M, Mazzoni MG, Raimondi M, Panigada M, Belliato M, Bronzini N, Latronico N, Petrucci N, Belgiorno N, Tagliabue P, Cortellazzi P, Gnesin P, Grosso P, Gritti P, Perazzo P, Severgnini P, Ruggeri P, Sebastiano P, Covello RD, Fernandez-Olmos R, Fumagalli R, Keim R, Rona R, Valsecchi R, Cattaneo S, Colombo S, Cirri S, Bonazzi S, Greco S, Muttini S, Langer T, Alaimo V, Viola U, (2020) Baseline Characteristics and Outcomes of 1591 Patients Infected With SARS-CoV-2 Admitted to ICUs of the Lombardy Region, Italy. JAMA 10.1001/jama.2020.5394

23. Wang D, Hu B, Hu C, Zhu F, Liu X, Zhang J, Wang B, Xiang H, Cheng Z, Xiong Y, Zhao Y, Li Y, Wang X, Peng Z, (2020) Clinical Characteristics of 138 Hospitalized Patients With 2019 Novel Coronavirus–Infected Pneumonia in Wuhan, China. JAMA 323: 1061–1069

24. Alhazzani W, Moller MH, Arabi YM, Loeb M, Gong MN, Fan E, Oczkowski S, Levy MM, Derde L, Dzierba A, Du B, Aboodi M, Wunsch H, Cecconi M, Koh Y, Chertow DS, Maitland K, Alshamsi F, Belley-Cote E, Greco M, Laundy M, Morgan JS, Kesecioglu J, McGeer A, Mermel L, Mammen MJ, Alexander PE, Arrington A, Centofanti JE, Citerio G, Baw B, Memish ZA, Hammond N, Hayden FG, Evans L, Rhodes A, (2020) Surviving Sepsis Campaign: Guidelines on the Management of Critically Ill Adults with Coronavirus Disease 2019 (COVID-19). Crit Care Med 48: e440–e469

25. Alhazzani W, Moller MH, Arabi YM, Loeb M, Gong MN, Fan E, Oczkowski S, Levy MM, Derde L, Dzierba A, Du B, Aboodi M, Wunsch H, Cecconi M, Koh Y, Chertow DS, Maitland K, Alshamsi F, Belley-Cote E, Greco M, Laundy M, Morgan JS, Kesecioglu J, McGeer A, Mermel L, Mammen MJ, Alexander PE, Arrington A, Centofanti JE, Citerio G, Baw B, Memish ZA, Hammond N, Hayden FG, Evans L, Rhodes A, (2020) Surviving Sepsis Campaign: guidelines on the management of critically ill adults with Coronavirus Disease 2019 (COVID-19). Intensive Care Med 46: 854–887

26. 26. Zhou F, Yu T, Du R, Fan G, Liu Y, Liu Z, Xiang J, Wang Y, Song B, Gu X, Guan L, Wei Y, Li H, Wu X, Xu J, Tu S, Zhang Y, Chen H, Cao B, (2020) Clinical course and risk factors for mortality of adult inpatients with COVID-19 in Wuhan, China: a retrospective cohort study. Lancet 395: 1054–1062

27. Ruan Q, Yang K, Wang W, Jiang L, Song J, (2020) Clinical predictors of mortality due to COVID-19 based on an analysis of data of 150 patients from Wuhan, China. Intensive Care Med 46: 846–848

28. Chan JF, Yuan S, Kok KH, To KK, Chu H, Yang J, Xing F, Liu J, Yip CC, Poon RW, Tsoi HW, Lo SK, Chan KH, Poon VK, Chan WM, Ip JD, Cai JP, Cheng VC, Chen H, Hui CK, Yuen KY, (2020) A familial cluster of pneumonia associated with the 2019 novel coronavirus indicating person-to-person transmission: a study of a family cluster. Lancet 395: 514–523

29. Helms J, Tacquard C, Severac F, Leonard-Lorant I, Ohana M, Delabranche X, Merdji H, Clere-Jehl R, Schenck M, Fagot Gandet F, Fafi-Kremer S, Castelain V, Schneider F, Grunebaum L, Angles-Cano E, Sattler L, Mertes PM, Meziani F, Group CT, (2020) High risk of thrombosis in patients with severe SARS-CoV-2 infection: a multicenter prospective cohort study. Intensive Care Med 10.1007/s00134-020-06062-x

30. Xu Z, Shi L, Wang Y, Zhang J, Huang L, Zhang C, Liu S, Zhao P, Liu H, Zhu L, Tai Y, Bai C, Gao T, Song J, Xia P, Dong J, Zhao J, Wang FS, (2020) Pathological findings of COVID-19 associated with acute respiratory distress syndrome. Lancet Respir Med 10.1016/S2213-2600(20)30076-X

31. Arabi YM, Fowler R, Hayden FG, (2020) Critical care management of adults with community-acquired severe respiratory viral infection. Intensive Care Med 46: 315–328

32. Mitja O, Clotet B, (2020) Use of antiviral drugs to reduce COVID-19 transmission. Lancet Glob Health 8: e639–e640

33. Wang Y, Zhang D, Du G, Du R, Zhao J, Jin Y, Fu S, Gao L, Cheng Z, Lu Q, Hu Y, Luo G, Wang K, Lu Y, Li H, Wang S, Ruan S, Yang C, Mei C, Wang Y, Ding D, Wu F, Tang X, Ye X, Ye Y, Liu B, Yang J, Yin W, Wang A, Fan G, Zhou F, Liu Z, Gu X, Xu J, Shang L, Zhang Y, Cao L, Guo T, Wan Y, Qin H, Jiang Y, Jaki T, Hayden FG, Horby PW, Cao B, Wang C, (2020) Remdesivir in adults with severe COVID-19: a randomised, double-blind, placebo-controlled, multicentre trial. Lancet 395: 1569–1578

34. Arabi YM, Mandourah Y, Al-Hameed F, Sindi AA, Almekhlafi GA, Hussein MA, Jose J, Pinto R, Al-Omari A, Kharaba A, Almotairi A, Al Khatib K, Alraddadi B, Shalhoub S, Abdulmomen A, Qushmaq I, Mady A, Solaiman O, Al-Aithan AM, Al-Raddadi R, Ragab A, Balkhy HH, Al Harthy A, Deeb AM, Al Mutairi H, Al-Dawood A, Merson L, Hayden FG, Fowler RA, Saudi Critical Care Trial G, (2018) Corticosteroid Therapy for Critically Ill Patients with Middle East Respiratory Syndrome. Am J Respir Crit Care Med 197: 757–767

35. Bouadma L, Lescure FX, Lucet JC, Yazdanpanah Y, Timsit JF, (2020) Severe SARS-CoV-2 infections: practical considerations and management strategy for intensivists. Intensive Care Med 46: 579–582

36. Russell CD, Millar JE, Baillie JK, (2020) Clinical evidence does not support corticosteroid treatment for 2019-nCoV lung injury. Lancet 395: 473–475

37. Shang L, Zhao J, Hu Y, Du R, Cao B, (2020) On the use of corticosteroids for 2019-nCoV pneumonia. Lancet 395: 683–684

38. Kupferschmidt K, (2020) A cheap steroid is the first drug shown to reduce death in COVID-19 patients. Science https://doi.org/10.1126/science.abd3683

39. Rice TW, Rubinson L, Uyeki TM, Vaughn FL, John BB, Miller RR, 3rd, Higgs E, Randolph AG, Smoot BE, Thompson BT, Network NA, (2012) Critical illness from 2009 pandemic influenza A virus and bacterial coinfection in the United States. Crit Care Med 40: 1487–1498

40. McCullers JA, (2013) Do specific virus-bacteria pairings drive clinical outcomes of pneumonia? Clin Microbiol Infect 19: 113–118

41. Rhodes A, Evans LE, Alhazzani W, Levy MM, Antonelli M, Ferrer R, Kumar A, Sevransky JE, Sprung CL, Nunnally ME, Rochwerg B, Rubenfeld GD, Angus DC, Annane D, Beale RJ, Bellinghan GJ, Bernard GR, Chiche JD, Coopersmith C, De Backer DP, French CJ, Fujishima S, Gerlach H, Hidalgo JL, Hollenberg SM, Jones AE, Karnad DR, Kleinpell RM, Koh Y, Lisboa TC, Machado FR, Marini JJ, Marshall JC, Mazuski JE, McIntyre LA, McLean AS, Mehta S, Moreno RP, Myburgh J, Navalesi P, Nishida O, Osborn TM, Perner A, Plunkett CM, Ranieri M, Schorr CA, Seckel MA, Seymour CW, Shieh L, Shukri KA, Simpson SQ, Singer M, Thompson BT, Townsend SR, Van der Poll T, Vincent JL, Wiersinga WJ, Zimmerman JL, Dellinger RP, (2017) Surviving Sepsis Campaign: International Guidelines for Management of Sepsis and Septic Shock: 2016. Intensive Care Med 43: 304–377

42. Rhodes A, Evans LE, Alhazzani W, Levy MM, Antonelli M, Ferrer R, Kumar A, Sevransky JE, Sprung CL, Nunnally ME, Rochwerg B, Rubenfeld GD, Angus DC, Annane D, Beale RJ, Bellinghan GJ, Bernard GR, Chiche JD, Coopersmith C, De Backer DP, French CJ, Fujishima S, Gerlach H, Hidalgo JL, Hollenberg SM, Jones AE, Karnad DR, Kleinpell RM, Koh Y, Lisboa TC, Machado FR, Marini JJ, Marshall JC, Mazuski JE, McIntyre LA, McLean AS, Mehta S, Moreno RP, Myburgh J, Navalesi P, Nishida O, Osborn TM, Perner A, Plunkett CM, Ranieri M, Schorr CA, Seckel MA, Seymour CW, Shieh L, Shukri KA, Simpson SQ, Singer M, Thompson BT, Townsend SR, Van der Poll T, Vincent JL, Wiersinga WJ, Zimmerman JL, Dellinger RP, (2017) Surviving Sepsis Campaign: International Guidelines for Management of Sepsis and Septic Shock: 2016. Crit Care Med 45: 486–552

## References

1. Huang C, Wang Y, Li X, et al. Clinical features of patients infected with 2019 novel coronavirus in Wuhan, China. Lancet. 2020;395(10223):497–506.

2. Liu J, Liu Y, Xiang P, et al. Neutrophil-to-Lymphocyte Ratio Predicts Severe Illness Patients with 2019 Novel Coronavirus in the Early Stage. medRxiv. 2020:2020.2002.2010.20021584.

3. Chen G, Wu D, Guo W, et al. Clinical and immunologic features in severe and moderate forms of Coronavirus Disease 2019. medRxiv. 2020:2020.2002.2016.20023903.

4. Liu Y, Sun W, Li J, et al. Clinical features and progression of acute respiratory distress syndrome in coronavirus disease 2019. medRxiv. 2020:2020.2002.2017.20024166.

5. Liu J, Li S, Liu J, et al. Longitudinal characteristics of lymphocyte responses and cytokine profiles in the peripheral blood of SARS-CoV-2 infected patients. medRxiv. 2020:2020.2002.2016.20023671.

6. Wan S, Yi Q, Fan S, Lv J, Zhang X, Guo L, Lang C, Xiao Q, Xiao K, Yi Z, Qiang M, Xiang J, Zhang B, Chen Y, (2020) Characteristics of lymphocyte subsets and cytokines in peripheral blood of 123 hospitalized patients with 2019 novel coronavirus pneumonia (NCP). medRxiv

7. Chen L, Liu HG, Liu W, et al. Analysis of clinical features of 29 patients with 2019 novel coronavirus pneumonia. Chinese journal of tuberculosis and respiratory diseases. 2020;43(0):E005.

8. Zhang JJ, Dong X, Cao YY, et al. Clinical characteristics of 140 patients infected with SARS-CoV-2 in Wuhan, China. Allergy 2020

9. Wan S, Xiang Y, Fang W, Zheng Y, Li B, Hu Y, Lang C, Huang D, Sun Q, Xiong Y, Huang X, Lv J, Luo Y, Shen L, Yang H, Huang G, Yang R, (2020) Clinical features and treatment of COVID-19 patients in northeast Chongqing. J Med Virol 10.1002/jmv.25783

10. Li K, Wu J, Wu F, et al. The Clinical and Chest CT Features Associated with Severe and Critical COVID-19 Pneumonia. Invest Radiol 2020

11. Zheng S, Fan J, Yu F, et al. Viral load dynamics and disease severity in patients infected with SARS-CoV-2 in Zhejiang province, China, January-March 2020: retrospective cohort study. BMJ 2020; 369: m1443.

12. Cao B, Wang Y, Wen D, et al. A Trial of Lopinavir-Ritonavir in Adults Hospitalized with Severe Covid-19. N Engl J Med 2020

13. Grasselli G, Zangrillo A, Zanella A, et al. Baseline Characteristics and Outcomes of 1591 Patients Infected With SARS-CoV-2 Admitted to ICUs of the Lombardy Region, Italy. JAMA 2020

